# Reception of Respectful Maternity Care and Their Determinants Among Postpartum Mothers During Institutional Childbirth in East Wollega Zone Hospitals, West Oromia, Ethiopia, 2026

**DOI:** 10.64898/2026.05.18.26353527

**Authors:** Tahir H. Ahmed, Sileshi G. Abeya, Eshetu E. Chaka

## Abstract

Respectful maternity care [RMC] comprises the primary components of high-quality maternal health services. Evidence on RMC levels and determinants in Ethiopia is still inadequate. This study aimed to examine the reception and its determinants among postnatal women in government hospitals in the East Wallaga Zone, West Oromia.

An institution-based cross-sectional study was conducted from June to October 2025, within seven days post-delivery. A structured questionnaire based on the WHO RMC tools was used. The total RMC score proved robust reliability [Cronbach’s α = 0.808] and was organized using the 75th-percentile threshold. Factor analysis revealed basic RMC dimensions, while logistic regression was used to identify predictors of a promising RMC experience.

This study presented that only 46.8% of postpartum mothers received adequate RMC, with significant gaps in care. The main deficiencies comprised poor provider self-introduction, failure to call women by name, and infrequent communication and consent practices. Three key RMC dimensions were identified: privacy and consent, explanation and permission, and respectful communication.

Using multivariate analysis, interpersonal caring practices were robust predictors of positive RMC experiences. Explaining procedures with possible events, maintaining privacy, obtaining consent, prompt responsiveness, provider self-introduction, and calling mothers by name were significantly associated factors. Sociodemographic and maternal reproductive factors were not significantly associated after adjusting for confounders.

Finally, fewer than half [46.6%] of mothers experienced adequate RMC, which indicated major gaps in woman-centered care. Improving respectful interpersonal communication, informed consent, and maintaining privacy should be prioritized to boost the quality of maternal healthcare in the study area.

## Introduction

Pregnancy and childbirth are traditionally recognized as transformative experiences that support families and communities worldwide. Pregnancy is a period of immense pleasure and extreme vulnerability for a woman. It is a period in her life when her relationship with her maternity care providers can empower or scar her emotionally. Respectful Maternity Care [RMC] has been recognized and accepted as a basic human right. It is defined as care that is organized and provided to all women in a manner that respects their dignity, privacy, and confidentiality, protects them from harm and mistreatment, and promotes informed choice and continuous support during labor and childbirth [1]. With global initiatives in women’s health care still planned primarily to reduce women’s morbidity and mortality, an important shift in perspective has emerged towards recognizing respect, dignity, and gender equality as integral features of health care services [1, 2].

Despite the established importance of dignified care, women in many parts of the world still experience negative and abusive care during institutionalized childbearing services. According to the available evidence, approximately 54.5% of women in the world experience some form of disrespect & abuse [D & A] in the course of maternity services [3]. Various studies in the Sub-Saharan region have established that the rate of D&A in women during institutional childbirth in the region is terrifyingly high, with a pooled prevalence of 44% D&A during childbirth, with community-based estimates around 63% and facility-based around 37% [4]. The different forms of D&A in this region of Africa have been reported to include physical and verbal abuse, procedures to which women have not consented, violation of confidentiality, and detention in facilities for failure to pay for services [5, 6]. The negative experiences have been reported to affect the confidence of mothers and discourage them from seeking healthcare services in the future, in addition to deliveries by unskilled birth attendants [7].

A complex interplay of personal, systemic, and facility-based factors shapes the experience of receiving RMC. Some studies have demonstrated that educational status is a critical factor in shaping maternal experiences and satisfaction with the care received. Mothers with higher educational status were found to be more aware and demanding of RMC than mothers with lower educational status [8]. Researchers have also acknowledged other obstetric factors as critical determinants of RMC. For example, births during daytime shifts and by cesarean section were associated with higher RMC scores than spontaneous vaginal births, during which mothers might be more prone to compromised care because of overworked healthcare providers [9, 10]. Facility-based obstacles, such as shortages of skilled healthcare workers, a lack of critical medical equipment, and overburdened services, were critical obstacles to providing RMC [11, 12, 13].

Apart from maternal demographic factors, other factors contributed to the experiences of women in birthing, including those at the provider level and health systems. According to studies conducted in Ethiopia and Pakistan, women’s willingness to seek maternity services in the future would largely depend on the care experiences they received in the past [14, 15]. However, evidence also indicated that having a companion positively impacted women’s perceptions of RMC. However, this practice was not well executed in many low-resource facilities due to space limitations, which might have contributed to negative experiences for women during childbirth and discouraged them from seeking maternity services in the future.

Systemic barriers also include inadequate provider training in RMC techniques. Studies examining the effectiveness of RMC interventions in countries such as Uganda and Kenya have found that these interventions may reduce the frequency of D&A during childbirth [16]. Moreover, gender and power issues in healthcare settings played a role in the patient-provider relationships, with some women feeling disempowered due to paternalistic tendencies of healthcare providers. In high-income countries like the US, disparities in person-centered maternity care have been found to exist, with minority groups being at higher risk for mistreatment during childbirth [17, 18].

The development of tools such as the Mothers on Respect [MOR] Index and the Person-Centered Maternity Care [PCMC] Scale has advanced research in this field by tracking women’s experiences across diverse contexts. Educational approaches for healthcare service providers, particularly midwives and nurses, have been recognized as important for enhancing RMC and culture change in maternity services globally [19].

Therefore, based on existing evidence, this study hypothesized that sociodemographic, obstetric, and health service-related factors are significantly associated with the reception of RMC among postpartum mothers. Explicitly, factors such as mother-centered care were expected to increase the likelihood of receiving RMC. Thus, this study aimed to assess how postpartum mothers feel about RMC, as well as what affected it during births at East Wallaga Zone hospitals [EWZH].

## Methods and Materials

### Study area

A facility-based quantitative cross-sectional study design was used to investigate respectful maternity care practices and their determinants during facility-based childbirth in all public hospitals in the East Wallaga Zone [EWZ], West Oromia, Ethiopia. In 2007, the EWZ had a population of 1,213,503, expected to increase to 1,892,276 by 2025 with an annual growth rate of 2.52%. This includes 296,095 women aged 15-49 years, making up 24.4% of the population. This gives an estimated 29,367 pregnancies/births annually, of which 11,659 [39.7%] occurred in hospitals, according to Ministry of Health data. There are five hospitals, one of which is specialized and one of which is a referral hospital, as well as health centers and posts, located in the zone and offer comprehensive services for pregnant women and newborns, including antenatal care, delivery/C-section, instrumental births, and postnatal care.

### Study design and period

A cross-sectional facility-based study was carried out in West Oromia, Ethiopia, to evaluate the reception of RMC during institutional deliveries. The study was conducted from June to October 2025. The study participants were postpartum women who had delivered in selected hospitals in the past seven days and were willing to participate. Women with severe complications and those who refused to participate were not included. Ethical standards, such as voluntary participation, verbal consent, confidentiality, and withdrawal, were strictly followed.

### Study population

#### Source population

All mothers who delivered their babies in the public hospitals of EWZ within the first week of their delivery, with respect to HCPs working in labor and delivery units of those hospitals during the study period.

### Study population

Mothers who delivered in the selected hospitals during the study period and who were available and eligible for interviews within their first week post-delivery

### Inclusion *and* Exclusion criteria

*Inclusion:* Women who gave birth in the selected hospitals during the study period and were clinically stable and able to respond [in their first week of post-delivery].

*Exclusion:* Mothers with severe complications or altered consciousness in the first week post-delivery, and/or those who refused the interviews.

### Sample size determination and sampling procedures

The minimum sample size needed to assess the magnitude of CRMC during childbirth was determined by the single population proportion formula ***n* = [Z^2^ p [1-p]/d^2^]** with the assumptions of **P** [proportion of CRC] = 66.95% [20], degree of precision [d] =4, 95% CI, design effect of 2 and 5% non-response rate. Based on information provided by the hospitals, 4418 deliveries were attended in the five hospitals between June 22 and December 21, 2021.

Based on the six-month average delivery flow to each selected hospital, **4418**/6 = 736 average deliveries/month. The sample size for each hospital [nj] was calculated as the total sample size [n] multiplied by the average six-month delivery flow for each selected hospital [nj], divided by the sum of the six-month average maternal delivery flows. i.e.

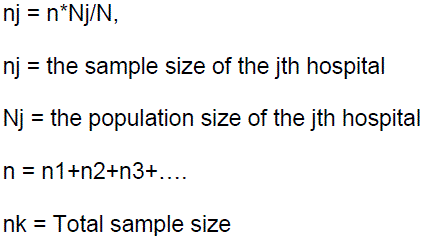

Based on these assumptions, a minimum of 618 + 5% contingence = 649 mothers participated in the study. For HCPs, all eligible and consenting mothers in the maternity/labor wards during the data-collection period were included.

“Sample size calculation was first done using Epi Info with an expected proportion of 36.95% from a previous study, requiring 618 participants for 95% confidence with 4% margin of error. However, the calculation did not include a finite-population correction, which is important given our population of 736 mothers.

In fact, we recruited all mothers who delivered at health facilities over 2 months, yielding a sample of 649. After removing 4 non-voluntary participants, 645 responses were included [87.6% of the target population].

With the actual proportion from our study [66.95%] [20], and taking into consideration the complex sampling design [design effect = 2.0] and the finite population correction, the precision reached is ±4.35% at 95% confidence level [95% CI: 62.60% – 71.30%]. This is actually better than what was originally intended since the proportion is actually farther from 50% than expected.

The sample comprised 645 mothers who gave birth in the selected hospitals and were interviewed for RMC and sociodemographic data. The sample size used in this study was determined to ensure sufficient precision to estimate RMC prevalence, EFA, and CFA.

### Data collection instruments and variables

A structured interview guide questionnaire consisting of four parts was used to assess mothers’ sociodemographic, obstetric, career-related conditions, and RMC domain. The RMC domain was sourced from WHO and publicly available RMC measurement tools, with items addressing privacy, dignity, and confidentiality; provision of information-sharing and informed consent; non-abuse practices; companion support, and maternal rights. It was asked using three-point response scales [i.e., “Yes, completely,” “Yes, to some extent,” “No, not at all”] and then coded for analysis. The questionnaire was developed in English, translated into Afan Oromo, and then back-translated to ensure concurrence of meaning [conceptual equivalence].

The structured questionnaire was adapted from the WHO RMC measurement tools and literature. The interviews were in quiet, private rooms to keep the comfort of the mothers. The questionnaire was tested in 5% [32] of the sample, adapted, and found to have face validity. Having always the correct technique of questionnaire administration during interviews, after practicing on motivation of participants, listening, and creating a good rapport with participants, refrain from asking leading questions and sensitive questions.

Responses were dichotomized by treating ‘Yes, completely’ and ‘Yes, to some extent’ as an indicator of RMC receipt, and ‘No, not at all’ as non-receipt of RMC. This method gives a conservative estimate of RMC, as women who responded ‘to some extent’ may still have had some gaps in care.

A composite Total RMC Score was created in SPSS version 25 as the dependent variable for regression analysis. It included the addition of 29 categorical variables [Q26-Q54] that assessed provider behavior during childbirth, including greetings, communication, autonomy, privacy, information exchange, and responsiveness, through Transform > Compute Variable with the formula Total_RMC_Score = SUM [Q26, Q27, …, Q54]. This dealt with missing data by requiring at least one non-missing value per case and provided a reliable scale [Cronbach’s α = 0.808] amenable to parametric analysis.

The Internal Consistency of the entire RMC scale and its individual components was examined by using Cronbach’s Alpha. A Cronbach’s Alpha of 0.70 or higher was considered acceptable for a multi-item instrument.

### Study variables

*Respectful Maternity Care [RMC]:* Organized care for all women, which respects their dignity, privacy, confidentiality, safety, absence of harm and mistreatment, and facilitates their informed decision-making and continuous support during labor and childbirth.

*Outcome variables*: Total RMC score [composite score of items] extracted from RMC factors [privacy & comfort; communication & information sharing; companion support & consent; maternal rights].

*Independent factors*:

1. *Sociodemographic characteristics* [age, sex, marital status, etc.]
2. *Reproductive health characteristics* [Age at first marriage, Age at first pregnancy, number of pregnancies, number of children born alive, number of abortions, number of stillbirths, ANC attendance, place of ANC, pregnancy status, mode of delivery]
3. *Dignity & Respect [Communication/Politeness]*: [HCP self-introduction, addressing mothers by name, Respectful/polite treatment, Language understood, treating with kindness/respect, answering Questions politely, speaking calmly/respectfully]
4. *Autonomy & Choice:* [Right to choose, invitation to ask questions, Companion invited to ask, respecting Choices/decisions, free choices of Companions, Allowing physical contact]
5. *Privacy & Confidentiality*: [Privacy, keeping Information secured]
6. *Communication & Information:* [Explanation of what to do, explaining labor events, explaining before actions, describing what you needed to do, explaining possible events
7. *Support & Responsiveness:* [Respecting mother’s beliefs, Respecting companion’s beliefs, considering mother’s comfort, Regular monitoring, telling to call for help, responding quickly to mother’s needs, caring/staying with mother *Adequate RMC:* the total RMC score ≥75th percentile.

### Data collectors and data collection procedures

Trained midwives and nurses carried out the data collection after a two-day training on the purpose of the study, ethics, interview techniques, questionnaire content, and data quality assurance. Interviews were conducted in a private facility setting during the first week after delivery, minimizing recall bias, with daily supervision by senior midwives to ensure completeness of the questionnaire. Pre-testing was conducted on 5% of the estimated sample size at Bako Hospital, followed by refinement. The data was double-entered for accuracy, and the coding of the data values was done by assigning “Yes, completely” a value of 2, “Yes to some extent” a value of 1, and “No, not at all” a value of 0. Scores for the components were calculated by making use of the regression method. The total and standardized scores were used for the analysis. The analysis of the multi-category data with latent normality was done by making use of the polychoric correlation method, while “minres” and “wls” with smoothing were used for the factor analysis for the resolution of non-normality, empty cells, and convergence problems. Translation/back-translation was used for the elimination of language bias. Tests used for psychometric validation included tests for the appropriateness of EFA/CFA by making use of KMO/Bartlett’s test, reliability by making use of Cronbach’s alpha, and lavaan and AMOS for the MR_Redcd_Amos.sav.

### Data management and analysis

The data were then entered into SPSS version 25. For data cleaning, range checks and checking for missing values were performed. For data description, the sociodemographic variables and the RMC item responses were summarized and presented in frequencies, percentages, means, and standard deviations, as appropriate. The suitability of the data for factor analysis was assessed. For the sampling adequacy sphericity tests, the Kaiser-Meyer-Olkin [KMO] and Bartlett’s test of sphericity were performed.

As regards factor extraction and rotation, exploratory and confirmatory factor analysis was performed using Principal Component Analysis [PCA] as factor extraction and Varimax rotation to simplify the component structure. The criteria for selecting the factors involved selecting those with eigenvalues of 1.0 and above, and interpreting the scree plot. Factors with loadings of 0.40 on each component were considered significant, and cross-loadings were resolved conceptually.

Bivariate tests [chi-square for categorical variables and t-tests or ANOVA for group comparisons of mean RMC score] were used to examine relationships between independent variables and RMC outcomes. Variables with p < 0.20 in bivariate tests were selected for multivariable modeling. Binary Logistic regression was used to identify independent predictors of RMC, controlling for potential confounding variables. Multicollinearity, predictor variables, and model fit were evaluated to declare statistical significance at p < 0.05.

## Data Preparation and Item Refinement

For item refinement, initial data screening was performed. Before the analysis, all negatively worded items were reverse-coded. Due to low item–total correlations, weak inter-item correlations, and poor factor loadings, several items were removed. Items related to treating women respectfully and politely, communicating peacefully, use of mothers’ language, and respect for mothers’ and companions’ beliefs were included.

A final set of 15 items was retained, demonstrating acceptable psychometric properties following iterative refinement. The retained items took key dimensions of RMC, including provider self-introduction, addressing mothers by name, autonomy in decision-making, encouragement to ask questions, privacy and confidentiality, clear explanations of care processes, regular monitoring, and responsiveness.

Warnings were observed due to unequal response distributions and the presence of sparse [zero] cells during estimation of the polychoric correlation matrix. These issues were considered expected due to the ordinal nature of the data, moderate sample size, and did not prevent further analysis.

### Ethical considerations

Wallaga University, Institute of Health Sciences Ethics Committee approved the Ethical clearance with Reference no. WU.SGS-58-2024. Permissions were obtained from the hospital administration. Participants’ consent to participate in the study was sought. Participants below 18 years old who participated in the study; their consent was processed according to government laws [their families were asked for their consent]. Participants had their privacy maintained by providing interview rooms for data collection. Participants’ identifiers were excluded from analysis. Participants had the right to withdraw from the study at any time.

### Quality control and limitations in methods

The quality of the data was assured through rigorous processes at all stages of data collection, entry, and validation. Trained midwives and nurses were given a two-day training session on the aims and objectives of the study, ethics, and interview and questionnaire content. In addition, there was daily supervision by senior midwives. Pre-testing of the questionnaire was done on 5% of the sample [32 mothers] at Bako Hospital. Double data entry to reduce the chances of errors, and since the data was in numbers [“Yes, completely” = 2, “Yes to some extent” = 1, “No, not at all” = 0], reverse scoring was used appropriately. The psychometric integrity of this study was ensured by using the KMO and Bartlett tests to check the suitability of the data for factor analysis, Cronbach’s alpha reliability test [α = 0.808], and model fit indices using lavaan and AMOS software for the MR_Redcd_Amos.sav file. The translation/back-translation method was used to eliminate linguistic biases, and reliable and representative data were obtained for RMC analysis

## Results

### 1. Sociodemographic characteristics of Postpartum women during institutional childbirth in EWZHs, West Oromia, Ethiopia

The mean age of postnatal mothers was 27.69 years, with a standard deviation [SD] of 4.968. Of the participants, 535 [82.9%] were between 20 and 34, followed by 85 [13.2%] who were older than 35 years. Regarding the marital status, the majority, 571 [88.5%], were married, and 36 [5.6%] were single. Almost half, 318 [49.3%] of the mothers were protestants, followed by 140 [21.7%] who were Orthodox. [see Figure 1]. The majority, 552 [85.58%], of the mothers were from the Oromo ethnic group, and 52 [8.07%] were Amhara. [See Table 1]

**Figure 1.**
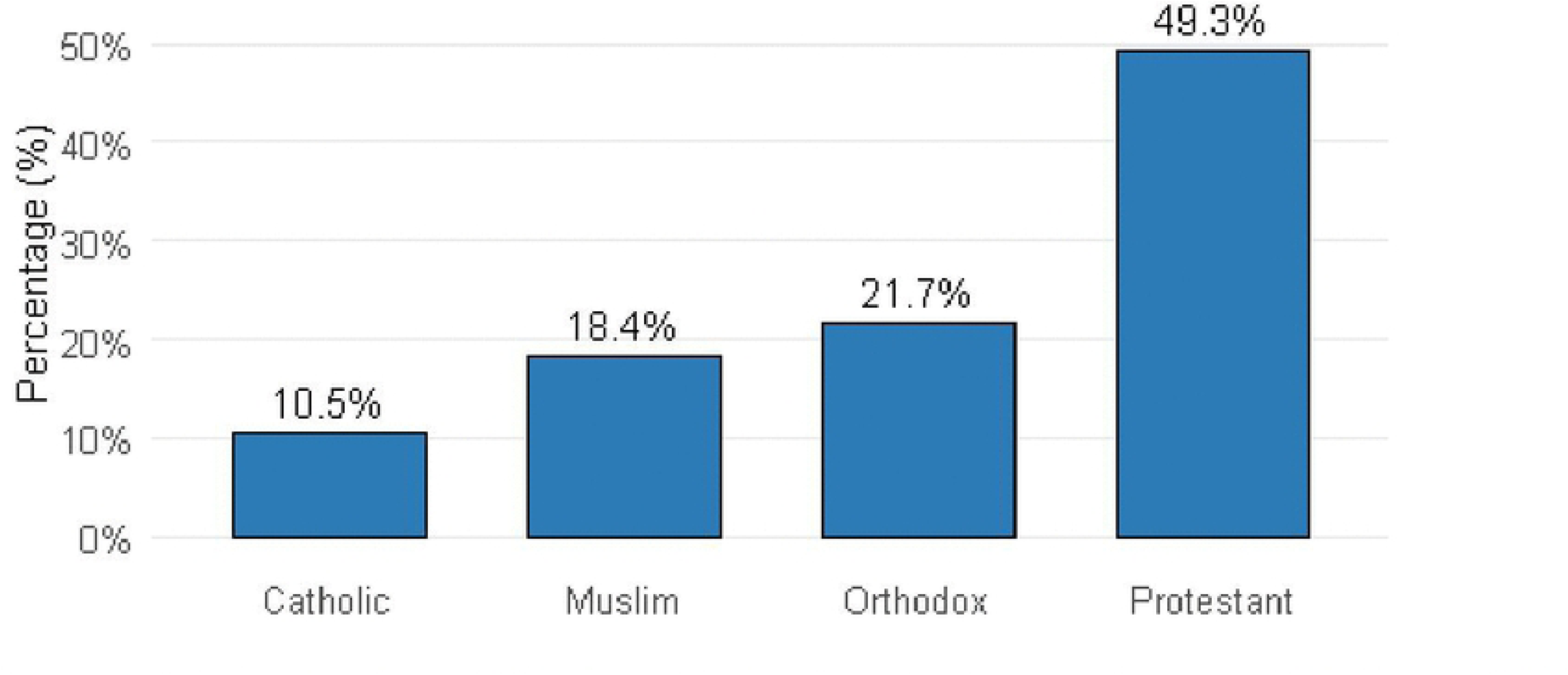
Distribution of Mothers’ Religion during Institutional Delivery in East Wallaga hospitals, West Oromia, Ethiopia, 2026.

**Table 1.** Sociodemographic Characteristics of women during institutional childbirth in EWZHs, West Oromia, Ethiopia, 2026.

| Variables | Categories | Frequency | Percent |
| --- | --- | --- | --- |
| **Marital status** | Married | 571 | 88.5 |
|  | Single | 36 | 5.6 |
|  | Divorced | 25 | 3.9 |
|  | Widowed | 13 | 2.0 |
| **Ethnicity** | Oromo | 552 | 85.6 |
|  | Amhara | 52 | 8.1 |
|  | Gurage | 27 | 4.2 |
|  | Tigre | 14 | 2.2 |
| **Occupational status** | Housewife | 377 | 58.4 |
|  | Government employee | 144 | 22.3 |
|  | Private employee | 81 | 12.6 |
|  | Student | 43 | 6.7 |
| **Monthly income** | <5000 | 262 | 40.6 |
|  | 5000-8000 | 275 | 42.6 |
|  | >8000 | 108 | 16.7 |

Regarding the educational status of the mothers, the majority, 264 [40.9%], were in grades 9-12, and college and above, 233 [36.18%]. [See Figure 2]

**Figure 2.**
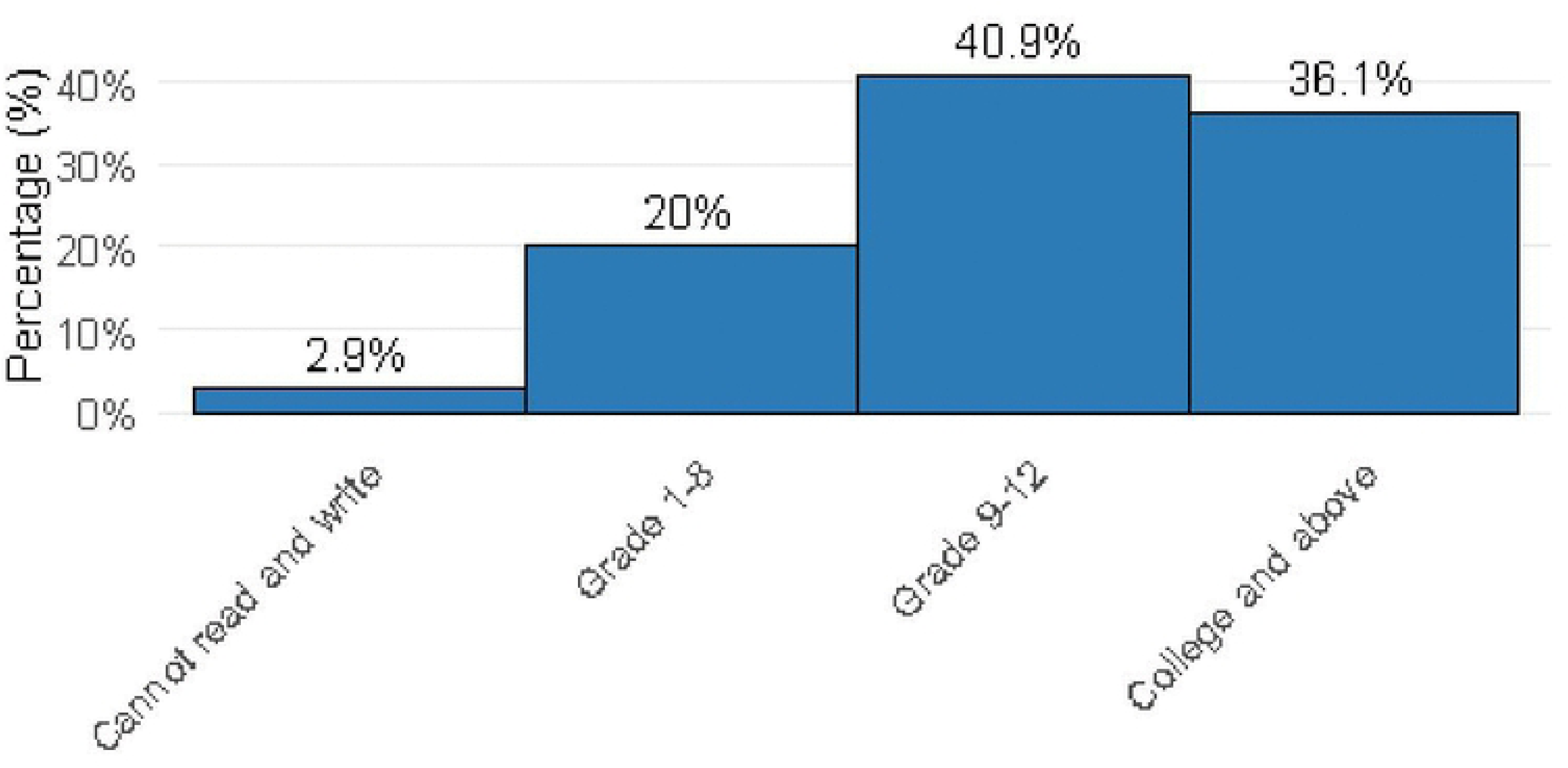
Distribution of Educational Status among Study Participants in West Oromia Hospitals, Ethiopia, 2026.

Regarding occupational status, housewives 377 [58.4%] were the majority, followed by 144 [22.3%] government employees, and 81 [12.6%] were private employees. The mean income of mothers was 6082.10 birr/month, with a standard deviation of 2504.38 birr/month. The majority, 275 [42.6%], earned between 5000 and 8000 birr, followed by 262 [40.6%], who earned less than 5000 birr, and a few, 108 [16.7%], earned more than 8000 birr/month. Almost two-thirds of the mothers were urban residents. [See figure 3]

**Figure 3.**
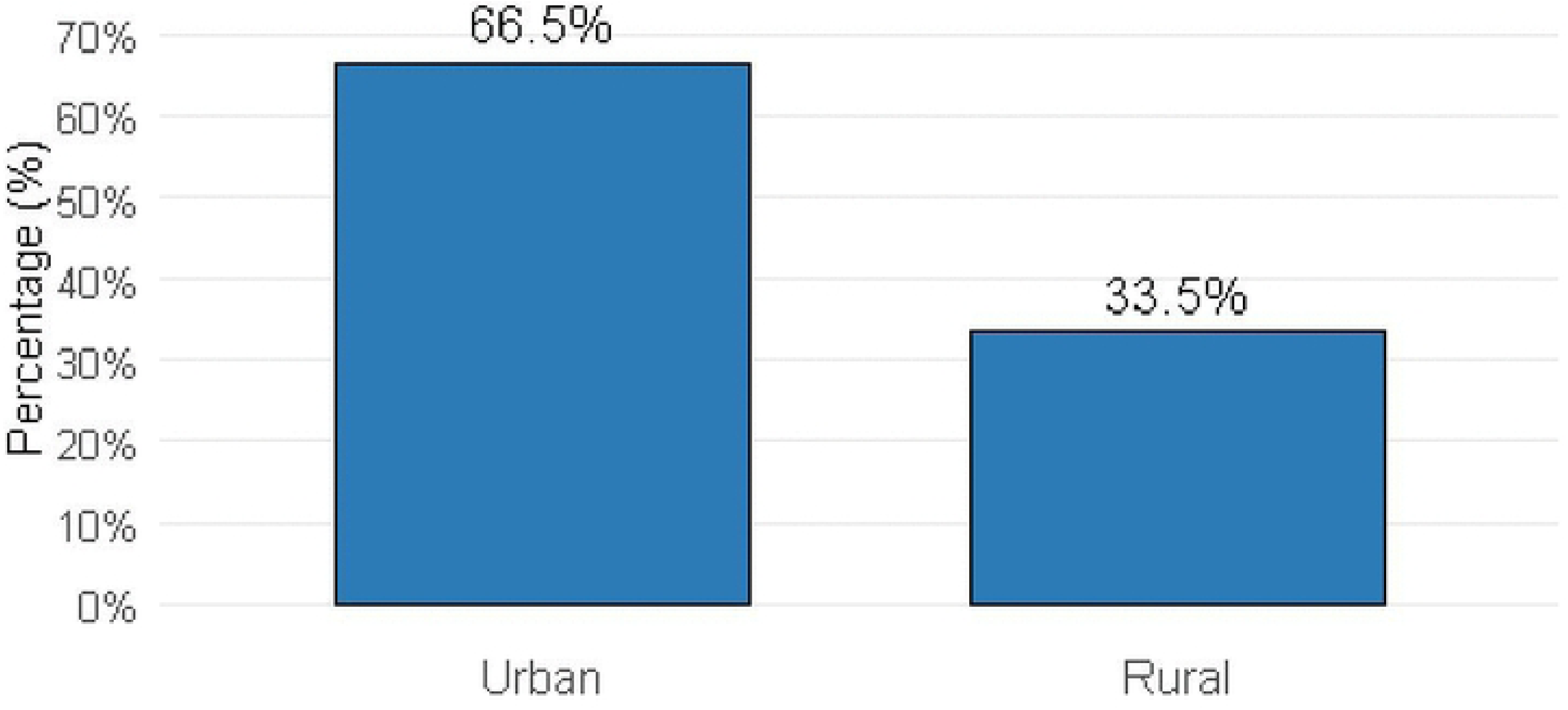
Frequency Distribution of Residence among Study Participants in West Oromia Hospitals, Ethiopia, 2026.

### 2. Reproductive Health Characteristics of women towards RMC and its determinants during institutional childbirth

The mean age = 21.56 at first marriage with a SD of 2.62. More than three-fourths, 504 [78.1%] of the women were married in the age range of 20 and 34 years, and 140 [21.7%] were married at 19 and less than 19 years. The mean age of the first pregnancy was 22.67 with a standard deviation of 2.643. The majority, 580 [89.9%], of the mothers got pregnant between 20 and 34 years old, and 61 [9.4%] became pregnant at age 19 or earlier. There were 2.72 pregnancies on average, with a SD of 1.5. Of the mothers, half, 328 [50.9%], had one or two children, and 227 [35.2%] had three or four children.

The average number of live births was 1.5 with a SD of 0.65. A third of the mothers had 216 [33.5%] three to four deliveries with live babies, whiles the majority, 370 [58.1%], had one to two deliveries and live babies. The average number of abortions among the mothers was 0.2 [SD = 0.414]. Most mothers, 526 [81.6%], had never experienced any abortion, while 114 [17.7%] reported only one abortion, and a very small number, 5 [0.8%], reported two. The mean number of miscarriages was 0.08 [SD = 0.283]. The majority of mothers had no history of stillbirth, 52 [8.1%] had experienced one stillbirth, and only 5 [0.2%] had experienced two. In terms of antenatal care [ANC] utilization, more than three-quarters of the mothers, 503 [78.0%], completed their ANC visits, while 131 [20.3%] attended ANC intermittently. [See table 2.]

**Table 2.** Reproductive Health Characteristics of women towards RMC and its determinants during institutional childbirth in EWZHs, West Oromia, Ethiopia 2026.

| Variables | Response Category | Frequency | Percent [%] |
| --- | --- | --- | --- |
| Age at first marriage | ≤19 | 140 | 21.7 |
|  | 20-34 | 504 | 78.1 |
|  | 35-50 | 1 | 0.2 |
| Age at first pregnancy | ≤19 | 61 | 9.4 |
|  | 20-34 | 580 | 89.9 |
|  | 35-50 | 4 | 0.7 |
| Number of pregnancies | 1-2 | 328 | 50.9 |
|  | 3-4 | 227 | 35.2 |
|  | 5-6 | 82 | 12.7 |
|  | ≥7 | 8 | 1.2 |
| Number of children born alive | 1-2 | 370 | 58.1 |
|  | 3-4 | 216 | 33.9 |
|  | 5-6 | 50 | 7.8 |
|  | ≥7 | 1 | 0.2 |
| Number of abortions | No abortion | 526 | 81.6 |
|  | 1 | 114 | 17.7 |
|  | 2 | 5 | 0.8 |
| Number of stillborn | baby born alive | 592 | 91.8 |
|  | 1 | 52 | 8.1 |
|  | 2 | 1 | 0.2 |
| Have attended ANC during this pregnancy | Yes completely | 503 | 78.0 |
|  | Yes, to some extent | 131 | 20.3 |
|  | No, not at all | 11 | 1.7 |
| Pregnancy status | Wanted and planned | 509 | 78.9 |
|  | Unwanted & unplanned | 136 | 21.1 |
| Birth attended by | Midwife | 378 | 58.6 |
|  | Doctor | 224 | 34.7 |
|  | Nurse | 14 | 2.2 |
|  | emergency surgery | 29 | 4.5 |
| The sex of the HCP mainly conducted labor | Male | 393 | 60.9 |
|  | Female | 252 | 39.1 |
| Birth complications | Yes | 229 | 35.5 |
|  | No | 416 | 64.5 |
| Types of maternal complications | Delayed second stage, CPD, prolonged labor, & Malpresentation & malposition. | 87 | 40.5 |
|  | Fetal Distress and IUFD. | 46 | 21.4 |
|  | Primary & Secondary PPH & First & second-degree perineal tears. | 44 | 20.5 |
|  | PIH and severe preeclampsia. | 28 | 13.0 |
|  | Infection | 10 | 4.7 |
| Instrumental delivery | Yes | 105 | 16.3 |
|  | No | 540 | 83.7 |
| Obstetric injury | First-degree tear | 18 | 2.8 |
|  | Second-degree tear | 6 | .9 |
|  | No injury | 621 | 96.3 |
| Outcome of labor | Healthy mother and newborn | 475 | 73.6 |
|  | Complicated maternal outcome | 67 | 10.4 |
|  | Fetal loss | 26 | 4.0 |
|  | Fetal complication | 77 | 11.9 |

### 3. Respectful Maternity Care for Postpartum Mothers during Institutional Childbirth

The study findings highlight numerous gaps in RMC, mainly in how healthcare providers [HCPs] communicate with mothers. Only about one-third of mothers, 215 [33.3%], stated that providers introduced themselves, whereas the majority, 430 [66.7%], said no such introduction was made. Likewise, nearly half of the mothers, 310 [48.1%], were never called by name during their care. About 187 [29.0%] said they were addressed by name only infrequently, and just 148 [22.9%] described being consistently called by their name. Finally, these findings recommend that care was often not personalized.

Mothers reported various experiences in terms of respectful and polite care. About one-third, 208 [32.2%], said they were treated with complete respect and politeness throughout their care, while 142 [22.0%] experienced this only to some extent. But a notable proportion, 295 [45.7%], reported that they did not receive respectful or polite treatment at all. Fascinatingly, a much larger share of mothers, 497 [77.1%], felt they were treated with kindness and respect overall. This proposes a difference between how mothers perceived specific polite behaviors and their wider sense of being treated kindly.

Many women still had limited participation in decisions about their care. Only 196 [30.4%] reported having full decision-making autonomy, while 199 [30.9%] had some level of involvement. However, a considerable proportion, 250 [38.8%], said they had no expressive role at all. Remarkably, despite these limitations, about three-quarters of mothers [479; 74.3%] felt that their treatment preferences were well respected. In addition, the freedom to choose a companion during care was even more widely supported, with 547 [84.7%] reporting that this choice was honored.

Encouragement for mothers and their companions to actively participate, particularly by asking questions, was commonly lacking. Only about one in four mothers [23.1%, 149] replied that HCPs felt completely motivated to ask questions, while a larger portion of mothers [39.4%, 254] reported receiving no encouragement at all. The situation was even more limited for companions, with nearly three-fourths [74.9%, 483] describing that they were not motivated to ask questions at all. In contrast, respect for individual beliefs and values was much more apparent. The majority of women [76.7%, 495] and an even larger number of companions [87.0%, 561] felt their beliefs were fully respected and supported, with very few reporting the absence of respect in this area.

Inclusive communication experiences were positive in many areas. Most women [86.2%, 556] entirely understood the language used by HCPs, although a smaller group 89 [13.8%] experienced partial or complete language barriers. In terms of responsiveness, more than half of the participants 360 [55.8%] reported that HCPs were reliably polite when responding to questions, while 252 [39.1%] labelled the responses as only partially polite. Likewise, most of the mothers 389 [60.3%] experienced calm and respectful communication. However, nearly one-third, 204 [31.6%], reported that they did not experience this level of respectful communication.

Physical contact, comfort, and privacy were generally well preserved. A large majority of postnatal mothers, 522 [80.9%], explained that permission was sought or that physical contact remained within acceptable parameters, while almost all 603 [93.5%] felt their comfort was accurately attended to. Privacy during examinations and delivery was also maintained for most women, with nearly 90% experiencing full or partial privacy, although around 10–12% stated a complete lack of privacy. On the contrary, the provision of information was uneven. Only 250 [38.8%] of women were wholly informed about data security and confidentiality, while nearly one-third, 187 [30%], got no information at all. Also, explanations about what to imagine during labor were often insufficient; just 193 [29.9%] conveyed being fully informed, while almost half, 308 [47.8%], received only incomplete explanations. In the meantime, 144 [22.3%] said they were not given any explanation, leading to unending gaps in informed and transparent care.

Monitoring of labor progress was inconsistent. Only 178 [27.6%] of postpartum women explained they were receiving complete and continuous monitoring, while a greater proportion, 253 [39.2%], said they were not supervised at all. Staff attendance during labor showed fairly better results, with 63.9% reporting consistent support; however, nearly one-quarter, 155 [24.0%], still experienced no staff presence during their labor and delivery.

Asking permission before performing procedures was inconsistent. About one-third of mothers 215 [33.3%] said that permission was not requested at all, while 245 [38.0%] replied that it was only partially sought. More than one-fourth, 185 [28.7%], designated that proper consent was reliably obtained. Furthermore, a substantial proportion of mothers 281 [43.6%] reported that their calls for help were not answered when needed. Despite this, responsiveness once needs were expressed was comparatively strong, with 559 [86.7%] of women receiving at least some form of timely response. Overall, nearly half, 302 [46.8%] of mothers received RMC. This recommends that more than half of the participants experienced care that did not meet RMC standards. [*See table 3*]

**Table 3.**
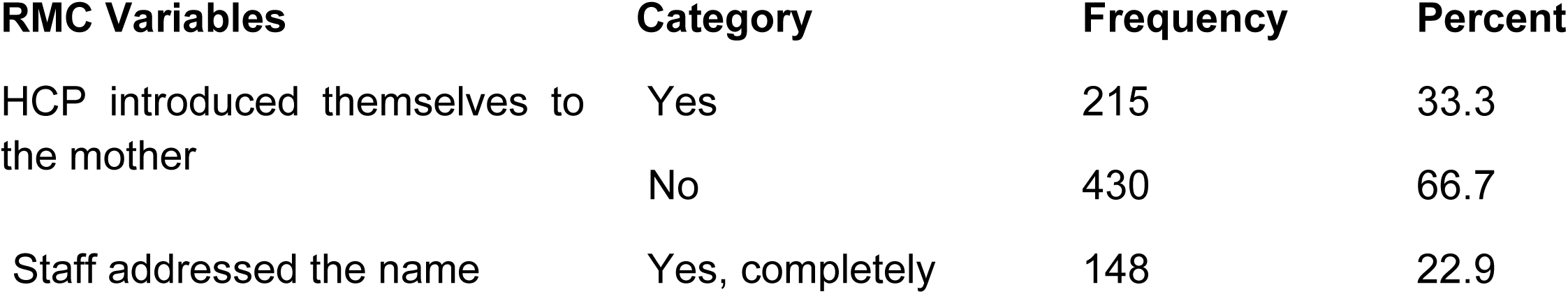
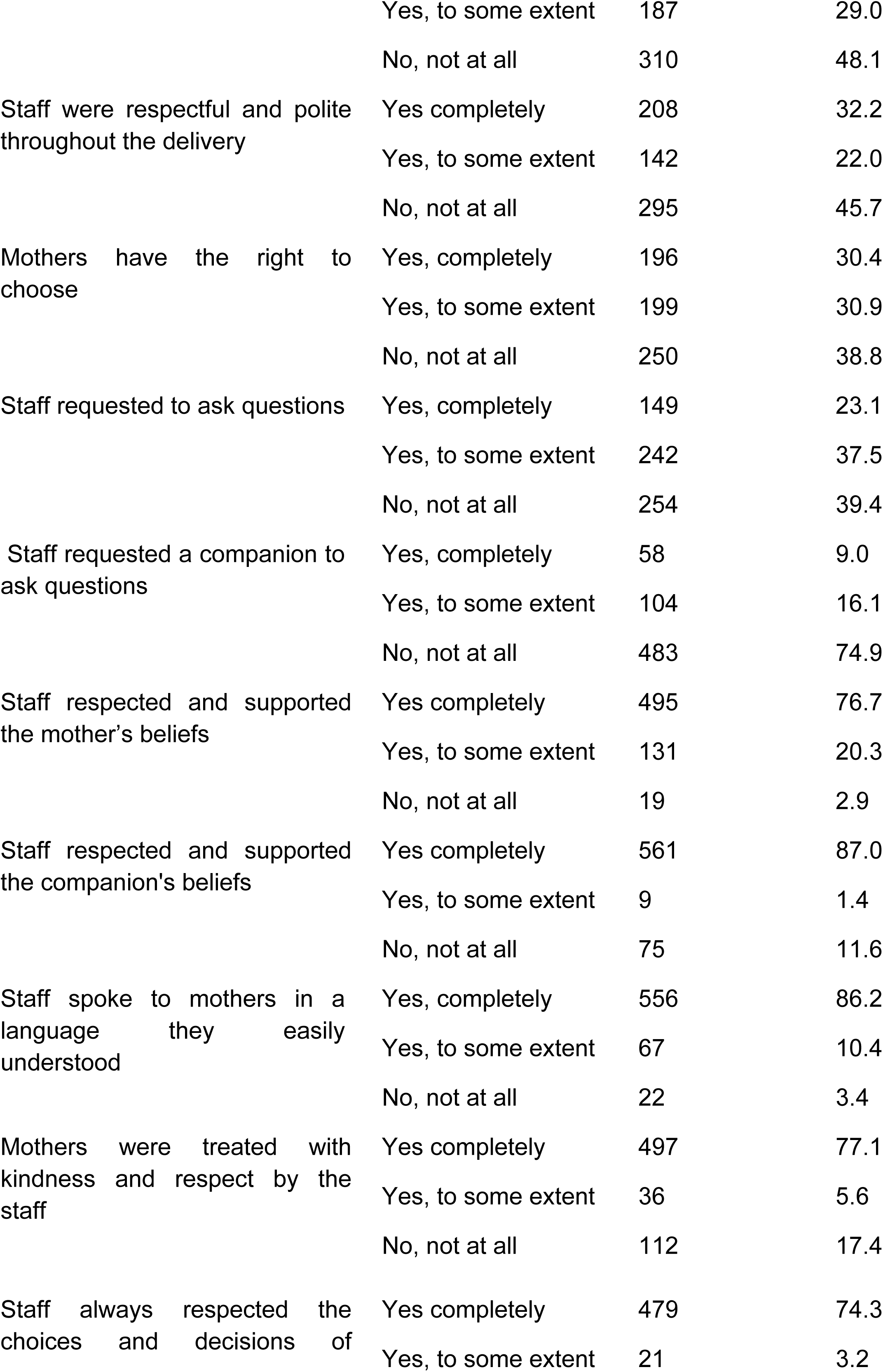
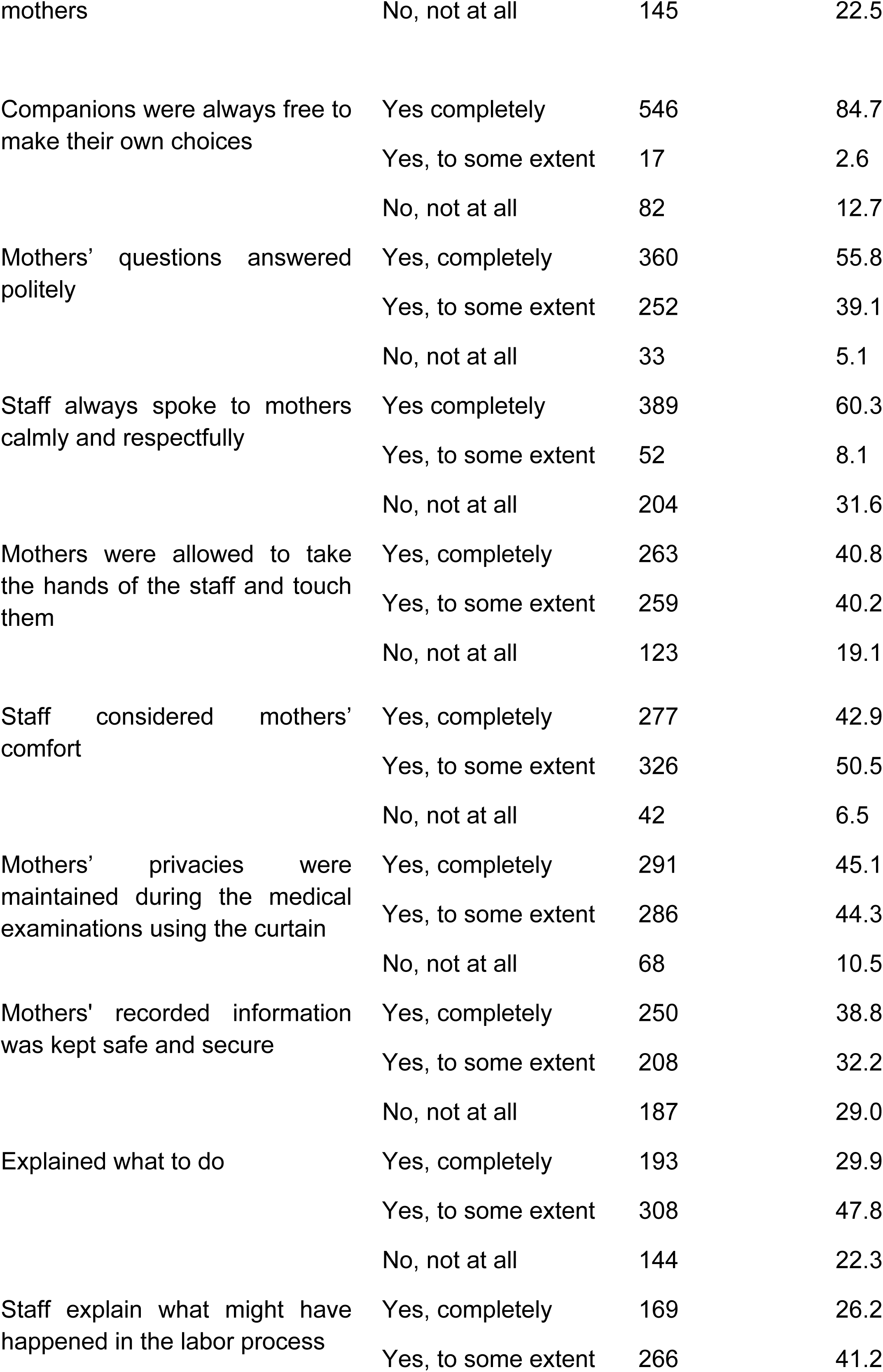
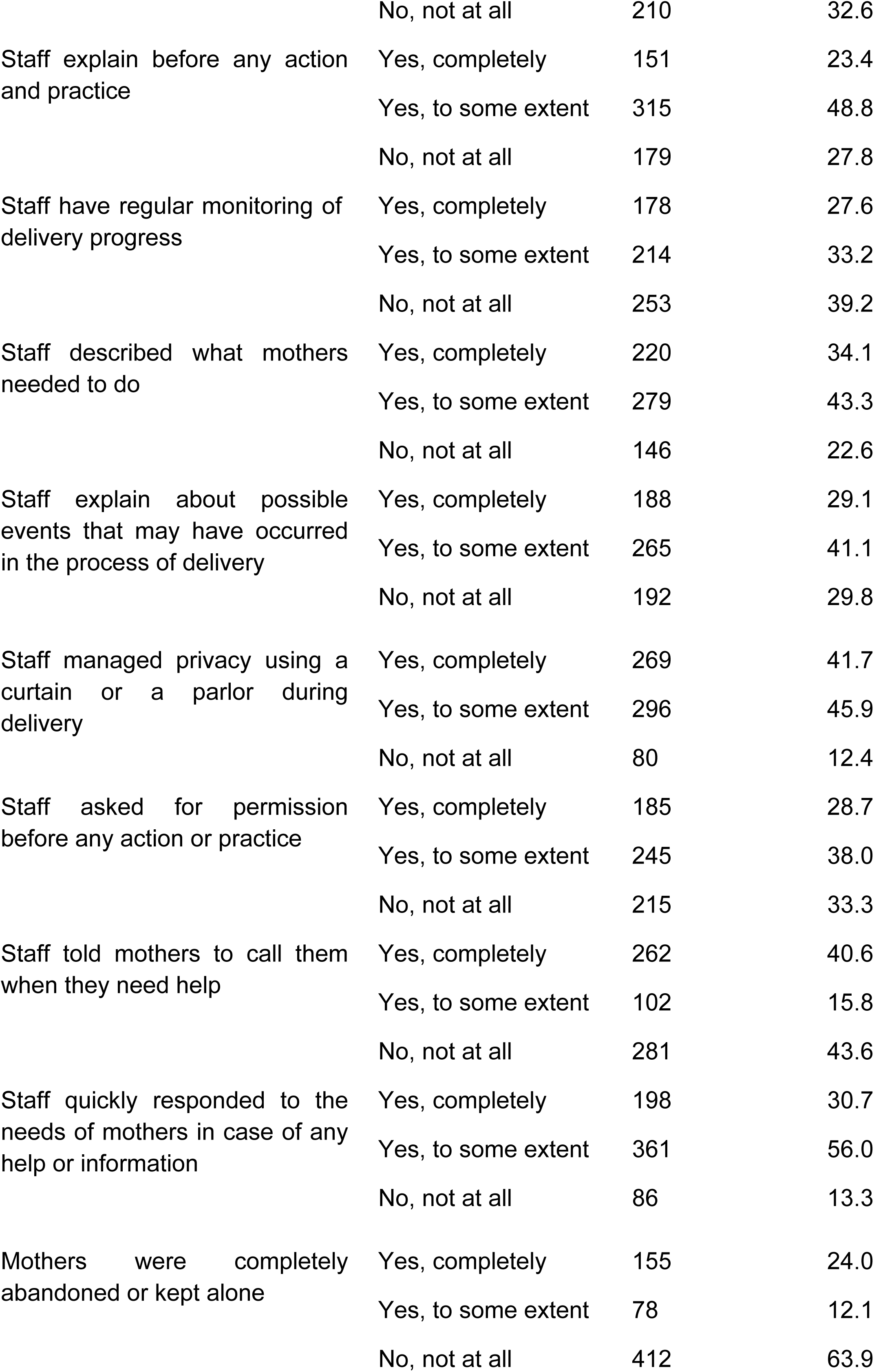
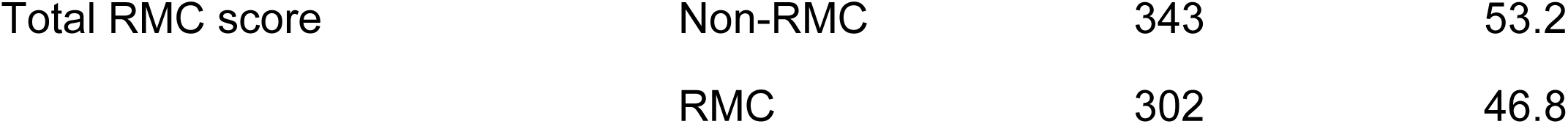
Respectful Maternity Care Practices and Their Determinants among Postpartum Mothers during Institutional Childbirth, in EWZHs, West Oromia, Ethiopia, 2026.

### 3. RMC-Specific Practices and RMC Status

Among the factors surveyed, RMC-related reception indicated the strongest association with overall RMC status. Healthcare provider self-introduction was particularly influential [χ² = 143.915, df = 1, p < .001], with 558 [86.5%] of the women receiving RMC compared to only 36.5% among HCPs who were not self-introduced. Other aspects, such as calling mothers by names, respecting their right to choose, encouraging them to ask questions, and explaining what to expect during labor, were also strongly associated with RMC status.

Other prominent factors included politeness in responses, appropriate physical touch and support, attention to comfort, maintenance of privacy, confidentiality of information, asking permission before procedures, responsiveness to women’s needs, and providing clear instructions, which were associated with RMC. On the contrary, factors such as overall kindness, respect for mothers’ choices, calm communication, and allowing freedom for companions showed relatively weaker associations with RMC status. [See Table 4]

**Table 4:** Association Between RMC Practices and RMC Status during institutional delivery in EWZHs, West Oromia, Ethiopia, 2026.

| Practice | $\chi^2$ [df] | p-value | Key Finding |
| --- | --- | --- | --- |
| HCP Self-Introduction | 143.915 | <0.001 | Strong association |
| HCP addressed women by Name | 80.421 | <0.001 | Higher RMC when complete |
| Right to Choose | 121.955 | <0.001 | Strong gradient effect |
| Encouraged Questions | 130.433 | <0.001 | Strong association |
| Explanation of Labor | 179.784 | <0.001 | Strongest predictor |
| Privacy | 65.127 | <0.001 | Strong association |
| Permission Before Action | 140.479 | <0.001 | Strong association |
| Quick Response | 130.443 | <0.001 | Strong association |
| Others |  | >0.05 | Weak or non-significant |

#### Independent Sample t-Test Findings

Independent sample t-tests were conducted and supported the findings from the chi-square analysis. Significant differences in mean RMC scores were observed across several factors, including religion [p < 0.001], income [p = 0.025], ANC follow-up [p < 0.001], place of ANC [p = 0.034], birth complications [p = 0.006], and mode of delivery. Notably, large differences were also found in main RMC practices such as HCP self-introduction [p < 0.001] and explanation of procedures [p < 0.001]. Remarkably, higher mean scores were observed in cases where certain RMC practices were absent.

In summary, numerous factors, including age, religion, occupation, parity, ANC attendance, mode of delivery, birth complications, labor outcomes, communication, privacy, informed consent, and responsiveness, revealed significant associations with RMC status. Among these, communication, privacy, informed consent, and responsiveness proved the strongest associations with RMC. But age, occupation, and complications during birth were not consistently associated with RMC status. Overall, the results recommend that key aspects of care, chiefly communication, respect for privacy, obtaining informed consent, and timely responsiveness, play a central role in improving the quality of maternal care.

#### Determinants of Positive RMC Experience [Main Model]

Multivariate logistic regression analysis was conducted to identify independent predictors of a positive RMC experience among postpartum women. Due to strong intercorrelations among the more than 25 RMC items surveyed in the univariate analysis, only seven key variables were carefully chosen for inclusion in the final model. These were selected based on their high Wald statistics, low p-values, and alignment with the WHO RMC domains, including effective communication, informed consent and autonomy, privacy and dignity, and continuous support.

The selected variables were: HCP self-introduction, addressing the mother by name, ensuring privacy using a curtain during examinations, explaining what to expect during labor, explaining procedures before performing them, obtaining permission before any action, and responding promptly to mothers’ needs or requests for help.

These seven key RMC variables, along with six socio-demographic and clinical confounders, age, number of pregnancies, age at first marriage, ANC attendance, place of ANC, and mode of delivery, were entered simultaneously into the multivariate model using the Enter method.

The overall model was strongly significant [Omnibus likelihood ratio test: χ² = 487.122, df = 21, p < 0.001], indicating that the predictors together provided a significantly better fit to the data than the null [intercept-only] model. The model established strong explanatory power, with a Nagelkerke pseudo-R² of 0.708 [Cox & Snell R² = 0.530], suggesting that nearly 70.8% of the variability in the outcome was explained by the included variables. Model convergence was attained normally [terminated at iteration 7 with parameter changes < 0.001].

Model calibration was excellent, as shown by the Hosmer–Lemeshow goodness-of-fit test [χ² = 4.653, df = 8, p = 0.794], which indicated no significant difference between observed and predicted event rates across the deciles of predicted probabilities. The contingency table further confirmed a close agreement between observed and expected frequencies across all risk groups, supporting good calibration of the model throughout the full range of predicted probabilities.

The model also confirmed strong discrimination performance. At the default probability cut-off of 0.500, it appropriately classified 86.5% of cases overall. The sensitivity for identifying positive RMC experiences was 86.0%, while the specificity for correctly identifying non-RMC cases was 87.1%.

After adjusting for socio-demographic and clinical variables, none of the confounders, including age, number of pregnancies, age at first marriage, ANC attendance, place of ANC, or mode of delivery, continued statistically significant [all p > 0.05]. In contrast, six of the seven RMC-related items emerged as strong and independent predictors of a positive RMC experience. [See, Table 5].

**Table 5:** Adjusted Odds Ratios [AOR] of Significant Respectful Maternity Care Predictors of Positive RMC Experience during institutional delivery in EWZHs, West Oromia, Ethiopia, 2026.

| Variables | Overall<br>Wald | df | p-value | AOR<br>[Exp[B]] | 95% CI<br>Lower | 95% CI<br>Upper |
| --- | --- | --- | --- | --- | --- | --- |
| Explanation of possible events in the labor process | 37.10 | 2 | <0.001 | 21.66 | 8.01 | 58.62 |
| Quick staff responsiveness to needs/help | 35.78 | 2 | <0.001 | 17.59 | 6.38 | 48.46 |
| Privacy was maintained during examinations using a curtain | 29.31 | 2 | <0.001 | 11.97 | 4.23 | 33.87 |
| Explanation before any action/practice | 18.82 | 2 | <0.001 | 7.32 | 2.65 | 20.21 |
| Permission was requested before any action/practice | 25.26 | 2 | <0.001 | 6.07 | 2.87 | 12.83 |
| The HCP introduced him/herself | 22.82 | 1 | <0.001 | 4.61 | 2.46 | 8.62 |
| Staff addressed the mother by name | 13.91 | 2 | 0.001 | 3.80 | 1.75 | 8.24 |

The strongest association was observed for explaining possible events during the labor process. Women who stated that staff “mostly” or “yes, completely” explained what to expect during labor had 21.66 times higher adjusted odds of experiencing positive RMC compared to those who reported “no, not at all” [AOR = 21.66, 95% CI: 8.01–58.62, p < 0.001]. The second strongest predictor was the quick responsiveness of staff to mothers’ needs or requests for help. Mothers who reported “mostly” or “yes” for responsiveness had 17.59 times higher adjusted odds of positive RMC experience [AOR = 17.59, 95% CI: 6.38–48.46, p < 0.001].

Maintaining privacy during medical examinations using a curtain was also a strong predictor of positive RMC experience. Mothers who replied “mostly” or “yes” for this practice had nearly 12 times higher adjusted odds [AOR = 11.97, 95% CI: 4.23–33.87, p < 0.001]. Providing explanations before any action or procedure was another significant factor, increasing the likelihood of positive RMC by more than sevenfold in the highest response category [AOR = 7.32, 95% CI: 2.65–20.21, p < 0.001]. In the same way, seeking permission before procedures was strongly associated with better outcomes, with “mostly/yes” responses linked to 6.07 times higher adjusted odds [AOR = 6.07, 95% CI: 2.87–12.83, p < 0.001]. HCP self-introduction was also associated with 4.61 times higher adjusted odds of positive RMC [95% CI: 2.46–8.62, p < 0.001]. Moreover, addressing mothers by name showed a meaningful association, with an adjusted odds ratio of 3.80 in the highest category [95% CI: 1.75–8.24, p = 0.001].

These findings suggested that, even after adjusting for potential confounders, key aspects of care, particularly effective communication [such as explaining labor events and procedures], responsiveness and support, maintenance of privacy, obtaining informed consent, and ensuring dignity and respect, were the main independent determinants of a positive RMC experience in this study.

#### RMC Subscale Analysis [Supplementary]

To complement the item-level analysis, a logistic regression model was fitted using aggregated RMC subscales [Respect, Privacy, and Care] derived from factor analysis. The model measured the predictive effect of these subscales on positive maternal outcomes [outcml_bin = 1], with 74% [n = 475] of respondents reporting positive outcomes. Generally, the model confirmed a good fit [Δ deviance = 18.52; AIC = 733.49].

Respect is the strongest predictor of positive outcomes. A one-point rise in the respect subscale was associated with higher odds of a positive outcome [OR = 2.22, 95% CI [1.43, 3.46], p < 0.001]. In contrast, Privacy showed a significant negative association, with a one-point increase linked to 46% lower odds of positive outcomes [OR = 0.54, 95% CI [0.36, 0.82], p = 0.004]. The Care subscale was not significantly associated with maternal outcomes [OR = 1.11, 95% CI [0.75, 1.65], p = 0.591]. **[See, Table 6.]**

**Table 6:**
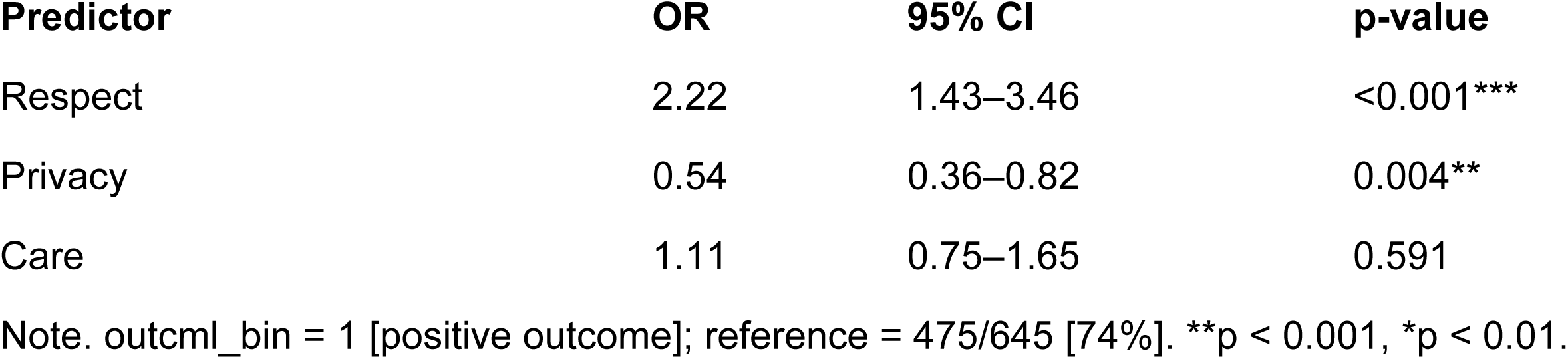
Logistic Regression: RMC Subscales Predicting Positive Maternal Outcomes [N = 645] during institutional childbirth in EWZHs, Oromia, Ethiopia, 2026.

These findings support the predictive validity of the RMC scale [EFA → CFA: CFI = 0.88; RMSEA = 0.10], with acceptable subscale reliability [e.g., Privacy α = 0.73]. Respect persisted as the strongest predictor of positive maternal outcomes, while covariates like age were not significant in supplementary models.

#### Predictive Validity: Main Effects and Interaction Testing Subscale Description

For regression analysis, the internal consistency and variability of the three RMC subscales were adequate. The highest mean score was the Respect dimension [mean = 2.58, SD = 0.41], followed by Privacy [mean = 1.83, SD = 0.55], while Care had the lowest mean [mean = 1.71, SD = 0.57]. [See, table 7]

**Table 7:** Descriptive and Reliability for RMC Subscales during institutional childbirth [N = 645] in EWZHs, Ethiopia, 2026.

| Subscale | Mean | SD | $\alpha$ |
| --- | --- | --- | --- |
| Respect | 2.58 | 0.41 | 0.52 |
| Privacy | 1.83 | 0.55 | 0.73 |
| Care | 1.71 | 0.57 | 0.59 |

**Table 8:**
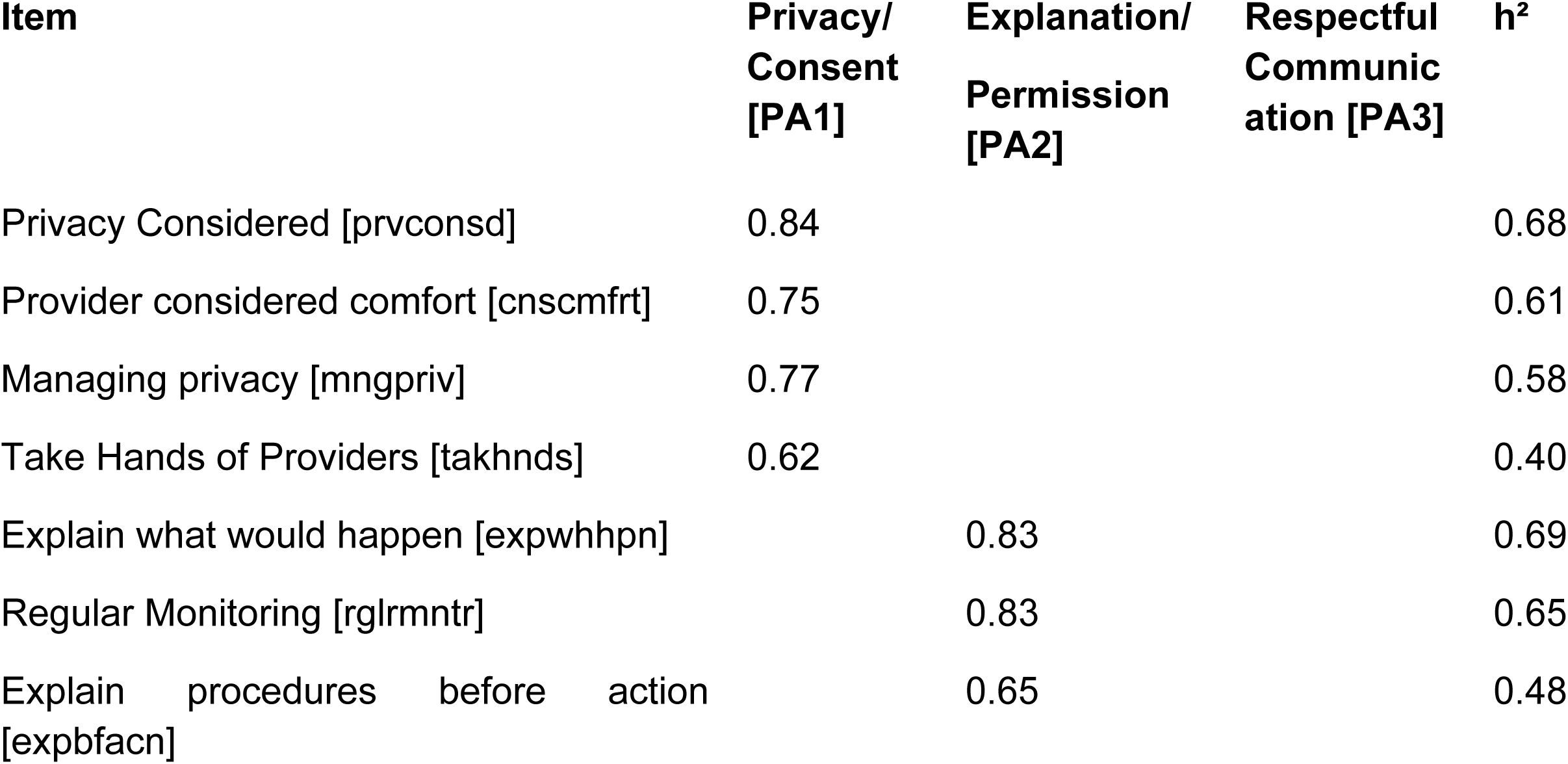
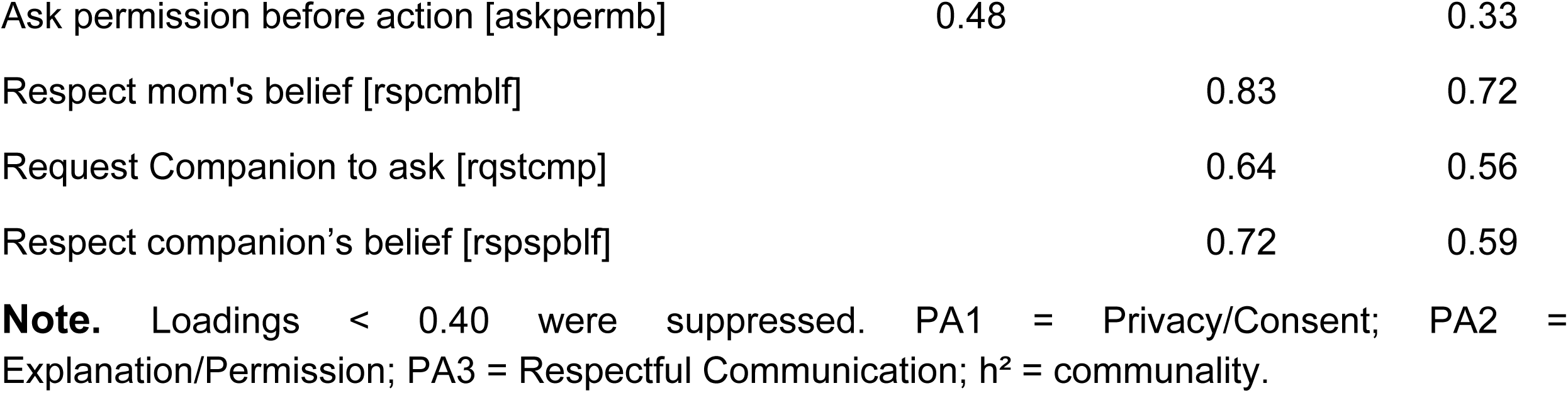
EFA Pattern Matrix Loadings [≥ .40] and Communalities during institutional delivery in EWZHs, West Oromia, Ethiopia, 2026, [n = 322].

**Table 9:** CFA Standardized Factor Loadings during institutional delivery in East Wallaga hospitals, West Oromia, Ethiopia,2026, [n = 323].

| Factor | Item | $\lambda$ | SE | z | p |
| --- | --- | --- | --- | --- | --- |
| Privacy | Mother's privacy considered [prvconsd] | 0.80 |  |  | < 0.001 |
|  | Mother's Comfort considered [cnscmfrt] | 0.66 | 0.070 | 11.84 | < 0.001 |
|  | Mothers take the hands of staff [takhnds] | 0.79 | 0.067 | 14.68 | < 0.001 |
| Explanation | Explained what has happened [expwhhpn] | 0.78 | 0.056 | 17.49 | < 0.001 |
|  | Explain before action [expbfacn] | 0.63 | 0.061 | 13.10 | < 0.001 |
|  | Staff ask permission from moms [askpermb] | 0.49 | 0.072 | 8.12 | < 0.001 |
| Respectful | Staff respected Companion belief [rspcmblf] | 0.66 | 0.065 | 12.35 | < 0.001 |
|  | Staff respected mom's belief [rspspblf] | 0.49 | 0.068 | 9.44 | < 0.001 |
|  | Staff spoke with the language moms understand [spklang] | -0.17 | 0.131 | -1.30 | 0.195 |

#### Respect × Privacy Interaction

Using a likelihood ratio test, the moderating effect of Respect and Privacy was assessed. The inclusion of the interaction term [Respect and Privacy] did not significantly improve model fit [χ²[1] = 2.57, p = 0.109], demonstrating no significant interaction between the two subscales. A hierarchical modeling approach [Multilevel Modeling] showed that the main effects-only model generated an adequate fit and was retained, as adding interaction terms did not produce a significant improvement based on χ² difference tests. Totally, the RMC scale validation process was finalized [EFA → CFA [CFI = 0.88; RMSEA = 0.10] → reliability → scoring → regression], supporting the use of the main effects model for subsequent analyses. [The validation process involved discovering the structure [EFA], testing it [CFA], and confirming reliability, creating scores, and then proving those scores to predict real outcomes].

#### Psychometric Evaluation of the Respectful Maternity Care [RMC] Scale

Using responses from mothers on a 3-point Likert-type scale [1 = “No, not at all,” 2 = “Yes, to some extent,” 3 = “Yes, completely”], the psychometric evaluation of the RMC scale was conducted. The dataset was randomly split into two subsamples: for exploratory factor analysis [EFA], 322 participants were used, and for confirmatory factor analysis [CFA], 323 participants were used. The analyses were performed in *R* [version 4.4.2] using the *psych* and *lavaan* packages. Due to the ordinal nature of the data, polychoric correlations were computed, and models were assessed using diagonally weighted least squares [DWLS] with strong standard errors.

#### Exploratory and Confirmatory Factor Analyses of the RMC Scale

The total sample was randomly split into two independent subsamples before conducting exploratory and confirmatory factor analyses. One subsample [n = 322] was used for EFA, while the other [n = 323] was used for CFA, in line with the recommended best practices in structural equation modeling.

#### Exploratory Factor Analysis

##### Assessment of Factorability

The suitability of the data was assessed before factor extraction. The Kaiser–Meyer–Olkin [KMO] measure designated adequate sampling adequacy [KMO = 0.83], with individual item values ranging from 0.57 to 0.93. Bartlett’s test of sphericity was significant [χ²[325] = 2422.04, p < 0.001], confirming that the correlation matrix was not an identity matrix and was appropriate for factor analysis.

##### Factor Extraction and Retention

A polychoric correlation matrix was used, given the ordinal nature of the RMC items. Using principal axis factoring [PAF], with oblimin rotation, factors were extracted. Three factors were retained based on eigenvalues and interpretability, with eigenvalues of 5.00, 4.40, and 2.48, explaining a total of 46% of the variance [19%, 17%, and 10%, respectively]. The mean item complexity was 1.4, and the RMSR was 0.06, demonstrating acceptable model fit. Factor correlations were weak to moderate [r = −0.14 to 0.24], supporting the use of oblique rotation and signifying related but distinct constructs. [See, Figure 7].

**Figure 4.**
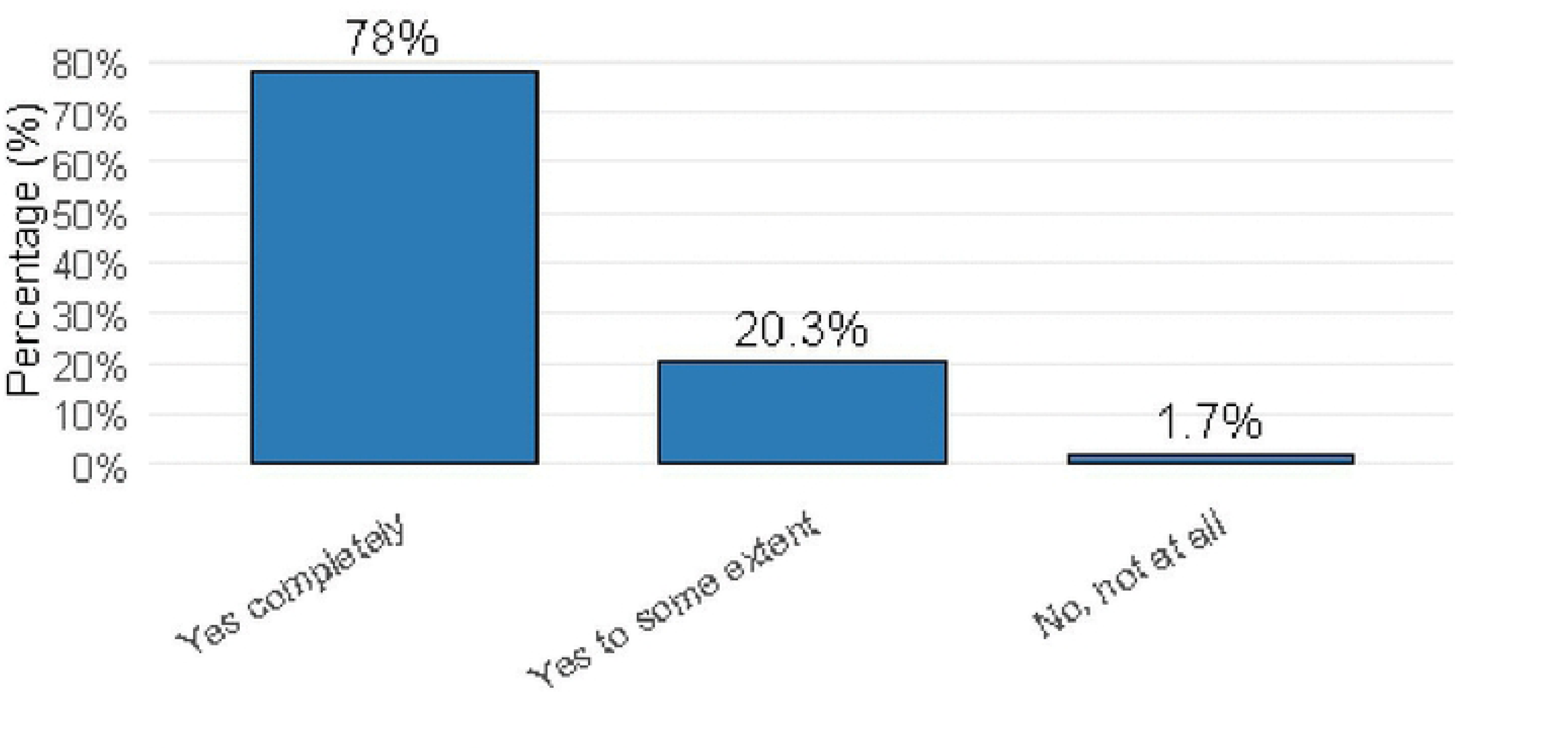
Frequency Distribution of ANC attendance among Study Participants in West Oromia Hospitals, Ethiopia,2026.

**Figure 5.**
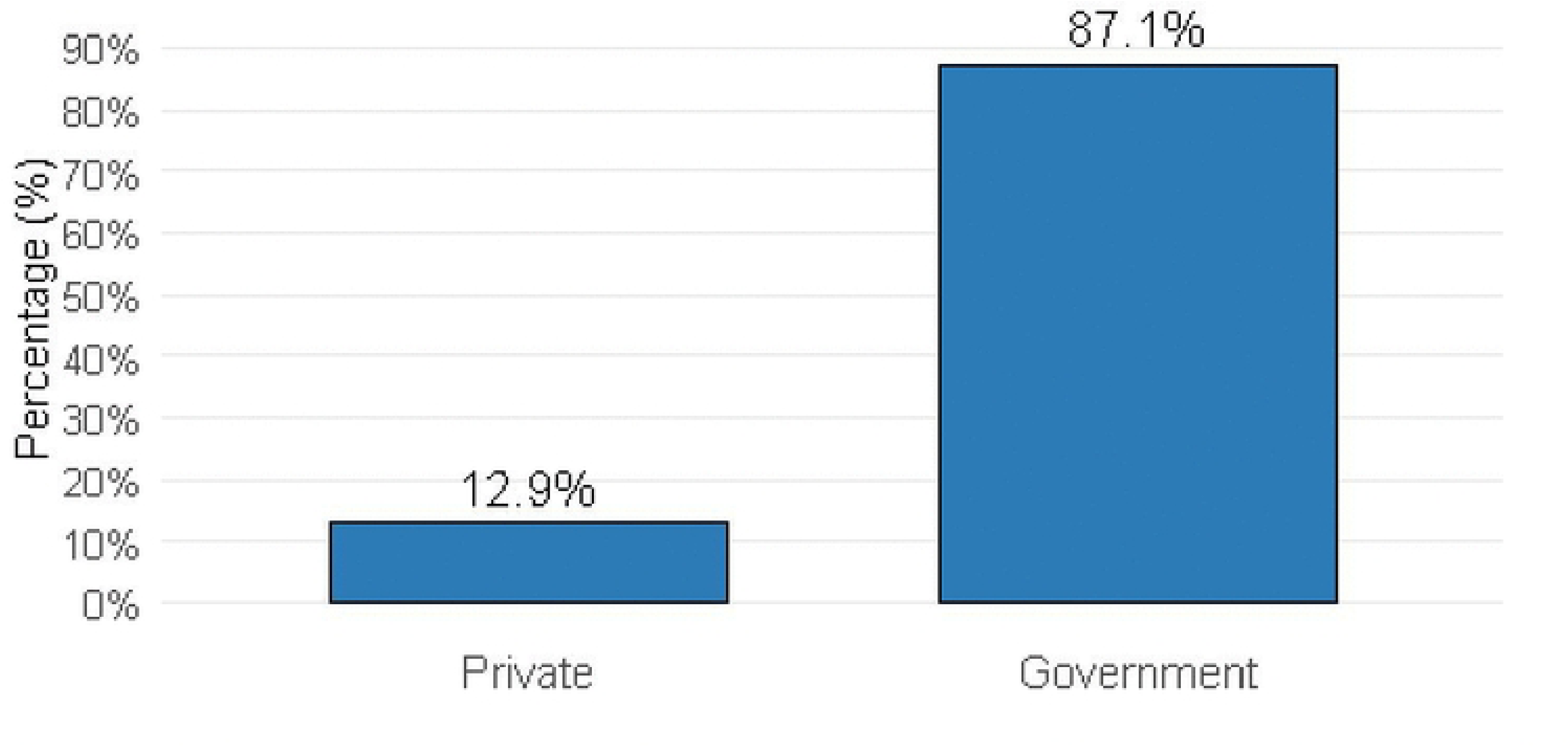
Frequency Distribution of Place of ANC among Study Participants in West Oromia EWZHs, Ethiopia, 2026.

**Figure 6.**
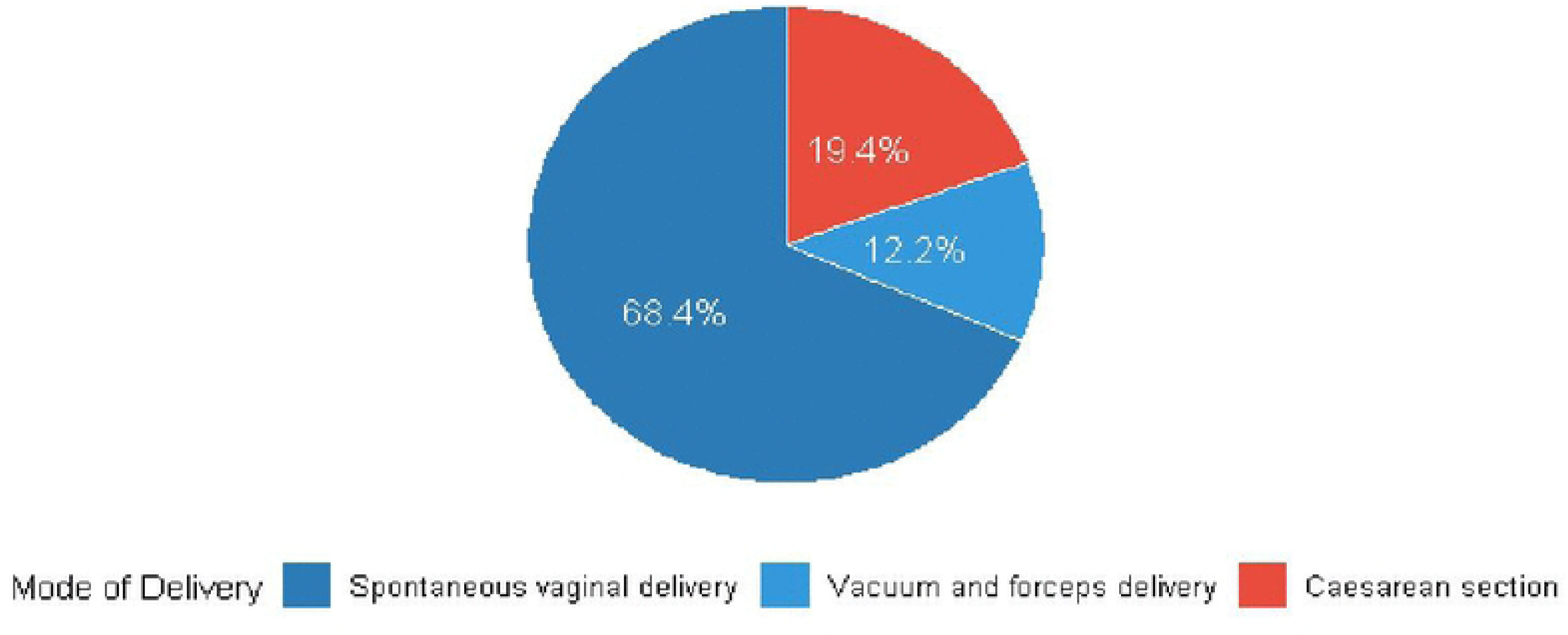
Frequency Distribution of Mode of Delivery among Study Participants in West Oromia EWZHs, Ethiopia, 2026.

### Factor Structure

Items with factor loadings ≥ 0.40 were retained. The three factors were classified as Privacy/Consent [Factor 1], Explanation/Permission [Factor 2], and Respectful Communication/Responsiveness [Factor 3]. All retained items proved acceptable communalities [≥ 0.30]. However, items such as *resplit*, *spklang*, *spkclmrs*, and *telcalnd* disclosed low communalities and were considered for exclusion during the refinement process. Generally, the EFA supported a theoretically coherent three-factor structure representing important domains of RMC.

### Confirmatory Factor Analysis

#### Model Specification

The three-factor structure identified in the EFA was further evaluated using confirmatory factor analysis [CFA] in an independent sample of 323 participants. The model was estimated using diagonally weighted least squares [DWLS], suitable for ordered categorical data. The CFA specified three correlated factors: Privacy/Consent, Explanation/Permission, and Respectful Communication/Responsiveness.

#### Model Fit

Model fit was assessed using multiple indices. The chi-square test was significant [χ²[51] = 315.37, p < 0.001]. Incremental fit indices showed marginal to acceptable fit [CFI = 0.905; TLI = 0.878]. But, absolute fit indices suggested poor model fit [RMSEA = 0.127, 90% CI [0.114, 0.141]; SRMR = 0.145], beyond suggested thresholds. Generally, the three-factor model demonstrated only partial fit.

#### Standardized Factor Loadings

Standardized factor loadings were usually moderate to strong and statistically significant [p < 0.01], except for spklang, which indicated a weak and non-significant loading [β = −0.17, p = 0.195]. Internal consistency was high for the Privacy [α = 0.85] and Explanation [α = 0.83] subscales, and adequate for Respectful Communication [α = 0.72].

### Factor Correlations

Privacy and Explanation were positively correlated [r = 0.53, p < 0.001]. Negative correlations were detected among Respectful Communication and Privacy [r = −0.26, p = 0.010] and between Respectful Communication and Explanation [r = −0.57, p < 0.001]. All correlations were below 0.85, supporting adequate discriminant validity, while the pattern suggests potential mis-specification of the Respectful Communication construct.

EFA supported a three-factor structure of the RMC scale [Privacy/Consent, Explanation/Permission, and Respectful Communication]. CFA results yielded partial support for this model, with tolerable incremental fit but poor absolute fit. The Privacy/Consent and Explanation/Permission factors showed stable performance, whereas the Respectful Communication factor performed less well, showing a need for further item refinement.

## Discussion

This study evaluated the scope of respectful maternity care [RMC], identified its fundamental dimensions, and examined its determinants among postpartum mothers in East Wallaga Zone hospitals. The findings exhibited that less than half [46.8%] of mothers received adequate RMC, demonstrating significant gaps in the quality of interactive and rights-based care during organizational childbirth. The analysis further proved that key domains of RMC—mainly Patient-centered care [communication, informed consent, privacy, and responsiveness] were the strongest predictors of optimistic maternal experiences, although sociodemographic factors were not independently associated with RMC after adjustment.

The overall magnitude of RMC observed in this study was lower than global recommendations that emphasized universal access to respectful and dignified maternity care [21]. However, this finding was consistent with previous studies conducted in low– and middle-income countries, including Ethiopia, where suboptimal levels of respectful care and experiences of D&A during childbirth had been widely reported [21, 22, 23]. Systematic and qualitative evidence have shown that women frequently experienced poor communication, lack of consent, and compromised dignity during facility-based childbirth [24, 25, 26]. The current study reinforced these findings and provided additional insight by using multidimensional measurement and statistical modeling to assess RMC.

One of the most significant findings of this study was the robust influence of communication-related practices on RMC. Mothers who got clear explanations of labor processes and procedures were significantly more likely to report optimistic care experiences. This finding was in line with earlier research indicating that effective communication augments trust, reduces anxiety, and improves overall maternal satisfaction [27, 28]. Likewise, the absence of informed consent and insufficient explanation of procedures had been identified as a major contributor to negative childbirth experiences universally [21, 29]. These findings suggested that healthcare providers might prioritize clinical tasks over patient-centered communication, thus discouraging the quality of care.

Responsiveness to mothers’ desires also became a key determinant of RMC. Women who described timely attention from HCPs meaningfully had higher odds of experiencing RMC. This was in line with studies conducted in comparable settings, where inattention, delays in care, and lack of provider availability were recognized as main contributors to dissatisfaction and poor maternal experiences [30]. Responsiveness mirrors both provider concentration and respect for women’s dignity, making it a vital component of quality maternity care.

Privacy and confidentiality were also found to be imperative factors persuading RMC. Mothers whose privacy was kept during examinations were more likely to report positive experiences. This finding is supported by previous studies, which have identified breaking of privacy as a common form of abuse that discourages women from using health facilities for childbirth [26, 31]. However, an unexpected result in this study was the negative association between the privacy subscale and positive outcomes. This may be described by increased awareness of privacy violations among dissatisfied mothers or potential measurement-related issues, such as reverse coding or perception bias. Similar encounters in measuring personal experiences of care have been reported in prior research [32].

The study also acknowledged gaps in basic respectful practices, such as HCPs introducing themselves and calling mothers by name. These simple yet essential aspects of RMC were strongly associated with positive outcomes. Previous studies have reported low levels of provider self-introduction and care personalization, contributing to feelings of neglect and depersonalization among mothers [26]. These results highlighted that refining RMC didn’t essentially need advanced resources but rather behavioral and cultural changes among HCPs.

In contrast, most sociodemographic and obstetric characteristics—with age, parity, and mode of delivery—were not significantly associated with RMC in the multivariable analysis. This suggested that RMC was chiefly influenced by HCP behavior and organizational practices rather than maternal characteristics. Though some studies have testified to associations between maternal characteristics and experiences of care [27], the present findings supported the principle that RMC should be generally provided to all women irrespective of their background. This has significant consequences for safeguarding equity in maternal healthcare.

The factor analysis showed in this study further reinforced the multidimensional nature of RMC, detecting key domains such as privacy/consent, explanation/permission, and respectful communication. These domains were consistent with existing frameworks and worldwide guidelines, which define RMC as encompassing dignity, autonomy, communication, and emotional support [28, 29]. However, the confirmatory factor analysis demonstrated only partial model fit; the findings largely supported the validity of the measurement style, yet represented the need for further improvement of RMC tools in comparable contexts.

From a policy and practice viewpoint, this study underscored the need for targeted interventions to advance communication, informed consent, and responsiveness in maternity care settings. Evidence recommended that training HCPs on RMC could meaningfully improve knowledge, attitudes, and skills [30, 33]. In addition, strengthening institutional policies, supportive supervision, and accountability mechanisms can help confirm adherence to RMC standards [23, 30]. Improving these aspects of care may improve maternal satisfaction, increase utilization of facility-based delivery services, and eventually contribute to improved maternal and neonatal outcomes.

This study has numerous strengths. It has used a relatively large sample size and applied robust statistical methods, including factor analysis and multivariable regression, to explore both the structure and determinants of RMC. The use of a multidimensional scale permitted an all-inclusive assessment of RMC practices.

Nevertheless, some restrictions should be recognized. The cross-sectional design limited the capability to establish causal relationships. The findings might also be affected by recall bias and societal desirability bias, although efforts were made to reduce these through early postpartum data collection and ensuring privacy during interviews. Moreover, the exclusion of mothers with severe problems may have led to an underestimation of negative care experiences. The partial model fit observed in factor analysis also suggested the need for additional proof of the RMC scale.

Future studies should consider longitudinal and mixed-methods approaches to better realize the causal pathways and contextual factors influencing RMC. Qualitative studies could offer bottomless insights into women’s lived experiences and HCPs’ perspectives, though interventional studies could evaluate the effectiveness of training and policy initiatives aimed at improving RMC.

In conclusion, this study revealed that RMC in this study area [EWZH] remained suboptimal, with noteworthy gaps in communication, informed consent, and responsiveness. These provider-related factors were the primary determinants of RMC, stressing the need for targeted interventions to advance the quality of maternal care. Strengthening RMC practices is indispensable for enhancing maternal experiences, endorsing equity, and refining overall health system performance.

## Strengths of the Study

- The study employed a large sample size [n = 645], ensuring adequate statistical power and precision.
- Use of validated WHO-adapted RMC tools enhanced content validity and comparability with global studies.
- Factor analysis [EFA and CFA] strengthened the psychometric validation of RMC dimensions.
- High internal consistency [Cronbach’s α = 0.808] confirmed the reliability of the composite RMC scale.
- Interviews conducted within seven days postpartum minimized recall bias.
- Inclusion of multiple public hospitals improved generalizability within the zone.
- Use of both descriptive and multivariable analyses provided a comprehensive determinant assessment.

## Limitations of the Study

- The cross-sectional design limited causal inference between determinants and RMC reception.
- Social desirability bias may have influenced mothers’ responses during facility-based interviews.
- Exclusion of severely ill mothers might have underestimated negative RMC experiences.
- The study relied on self-reported perceptions rather than direct observation of provider behavior.
- Private health facilities were not included, limiting comparative analysis across sectors.
- Cultural and provider-level factors [e.g., workload, training level] were not deeply explored.

### Recommendations

Based on the results of the study, the following were recommended: Providing regular training for healthcare providers in the area of RMC and patient communication. The need for the institutionalization of mandatory protocols for self-introduction by providers, informed consent, and explanation was also recommended by the study. To prepare mothers for their rights and expectations in childbirth, strengthening ANC counseling is a priority. Development of RMC monitoring and accountability frameworks at the hospital level is another priority. Improving staffing and workload management in maternity settings is a priority in order to improve RMC, which can result in better communication between providers and mothers, creating a more supportive environment for mothers during childbirth. Promotion of companion-inclusive maternity policies is another priority. Integration of RMC indicators into national maternal health quality improvement programs is a priority. Conducting longitudinal and mixed-method studies of the causal and experiential aspects of RMC was recommended.

## Conclusions

This study established that the experience of respectful maternity care among postpartum mothers in East Wallaga Zone hospitals was suboptimal, whereby more than half [53.2%] of the mothers experienced non-respectful care in institutional childbirth. While the experience of kind care, language comprehension, and respect for beliefs was satisfactory, gaps were observed in communication, informed consent, autonomy, and provider-mother interaction. Factor analysis revealed that RMC is a multidimensional construct comprising privacy/comfort, communication/information sharing, support/consent from companions, and mothers’ rights. Sociodemographic factors such as age, religion, and occupation, and obstetric factors such as parity, attendance at ANC sessions, mode of delivery, complications during delivery, and labor outcomes were significant predictors of RMC receipt. However, the most significant predictors of RMC reception in public hospitals in West Oromia, Ethiopia, were characteristics such as the self-introduction of care providers to mothers, explanation of procedures to mothers, provision of privacy, requesting consent before actions, and timely provision of responses to mothers’ needs. The findings revealed the need to address the issue of RMC reception with utmost urgency from an institutional level, provider level, and policy level to enhance the quality of RMC and mothers’ experiences during childbirth in public hospitals in West Oromia, Ethiopia.

## Data Availability

All related data are within the manuscript and its backup information files.

